# Subperiosteal hematoma of the ilium: a characteristic traction injury pattern causing acute groin pain in adolescent athletes

**DOI:** 10.64898/2026.02.06.26345690

**Authors:** Shinsuke Sakoda

**Author notes:** **Corresponding Author** Shinsuke Sakoda, MD, Department of Sports Medicine, Ashiya Central Hospital, 283-7 Yamaga, Ashiya-machi, Onga-gun, Fukuoka 807-0141, Japan. **Funding** This research received no external funding. **Ethical Approval** This study was approved by the institutional ethics committee of Ashiya Central Hospital and conducted in accordance with the Declaration of Helsinki. **Informed Consent** Informed consent was obtained using an opt-out method approved by the ethics committee due to the retrospective nature of the study.

## Abstract

**Purpose:** Subperiosteal hematoma of the ilium is an underrecognized traction injury causing acute groin pain in adolescent athletes and is often occult on plain radiographs. This study aimed to clarify the imaging features, clinical presentation, injury mechanisms, and return-to-sport course in a consecutively evaluated cohort.

**Methods:** This retrospective case series included adolescent athletes diagnosed at a single sports orthopaedic center between 2015 and 2025. Among 164 consecutive patients aged 22 years or younger presenting with acute sports-related groin pain and evaluated using a standardized MRI workflow, 20 met predefined MRI criteria. Demographics, skeletal maturity, sport, injury mechanism, ability to continue activity, treatment, and time to return to sport were analyzed descriptively.

**Results:** Mean age was 13.9 (SD 1.5) years; 95% were male and 90% had open physes. Soccer was the most common sport (65%). Injury mechanisms included kicking (50%), sprinting (20%), sudden deceleration (15%), and player contact (15%). Eighty-five percent were unable to continue sports immediately after injury. All patients were treated conservatively and returned to sport at a mean of 4.5 (SD 2.2) weeks. This condition accounted for 12.2% of adolescents evaluated with MRI for radiograph-negative acute groin pain in this cohort.

**Conclusion:** Subperiosteal hematoma of the ilium represents a reproducible traction injury pattern in skeletally immature athletes, particularly male soccer players, associated with kicking and sprinting. Awareness of this radiograph-occult but MRI-detectable entity may improve diagnostic accuracy and guide appropriate management of acute groin pain in adolescents.

## Introduction

Acute sports-related groin pain in adolescent athletes often presents a diagnostic challenge, particularly when plain radiographs reveal no apparent abnormalities. In routine clinical practice, the diagnostic approach to acute groin pain typically focuses on muscle strain, tendon injury, apophyseal avulsion, stress fracture, or sports hernia [1–5]. However, certain traction injuries around the iliac fossa are not routinely considered when radiographic findings are normal.

Subperiosteal hematoma of the ilium should be recognized as one such underrecognized traction injury. This condition involves periosteal stripping from the cortical surface at the iliac fossa, the attachment site of the iliacus muscle, with subsequent accumulation of fluid within the created subperiosteal space [6–10]. It has been reported to occur predominantly in adolescent athletes [6, 9].

During the growth period, the cortical bone is relatively thin while the periosteum remains comparatively strong, allowing traction forces generated by powerful muscle contractions to concentrate at the bone–periosteum interface [10–13].

In clinical settings, this condition is not always clearly differentiated from other causes of groin pain and may be managed as a nonspecific soft tissue injury, particularly when radiographic findings are unremarkable [3–5].

With advances in imaging modalities, MRI can readily demonstrate subperiosteal fluid collection, periosteal elevation, and surrounding reactive changes [4–6, 8]. This suggests that the condition may not be as rare as previously thought but rather underrecognized due to the limitations of initial plain radiographs.

The consistent association with kicking motion observed in this cohort provides a biomechanical explanation linking forceful iliacus contraction to periosteal stripping at the iliac fossa. This characteristic mechanism distinguishes subperiosteal hematoma of the ilium from other causes of acute groin pain and supports its recognition as a specific traction injury pattern during skeletal immaturity [1, 10–13].

The purpose of this study was to retrospectively analyze consecutive adolescent athletes diagnosed with subperiosteal hematoma of the ilium in order to clarify the imaging characteristics, clinical presentation, injury mechanisms, and return-to-sport course of this specific traction injury pattern.

## Methods

### Study Design

This study was a retrospective case series conducted at a single sports orthopaedic center between January 2015 and November 2025. The study was performed in accordance with the Declaration of Helsinki.

### Participants

During the study period, 164 consecutive patients aged ≤22 years who presented with acute sports-related groin pain were evaluated. Among these patients, 20 were diagnosed with subperiosteal hematoma of the ilium and were included in the final analysis. Exclusion criteria were evident high-energy trauma, the presence of a bleeding disorder, and postoperative hematoma.

### MRI protocol and definition of the denominator

At our institution, adolescent athletes presenting with acute sports-related groin pain undergo a standardized diagnostic workflow. When initial radiographs are unremarkable or do not adequately explain symptoms, MRI is routinely obtained to evaluate radiographically occult conditions.

Therefore, the denominator in this study (164 consecutive patients aged ≤22 years evaluated for acute sports-related groin pain during the study period) represents a consistently assessed clinical cohort under a uniform institutional practice, rather than a population-based sample.

Accordingly, the observed proportion of subperiosteal hematoma of the ilium should be interpreted as the relative frequency within this clinically evaluated cohort and not as the incidence in the general athlete population.

### Diagnostic Criteria for Subperiosteal Hematoma of the Ilium

The diagnosis of subperiosteal hematoma of the ilium was based strictly on predefined MRI criteria, including subperiosteal fluid collection along the iliac fossa, periosteal elevation, and/or reactive changes in the surrounding soft tissues, with exclusion of traumatic fractures and neoplastic lesions. MRI assessments were performed independently by two specialists: a sports medicine orthopaedic surgeon and a radiologist. In cases of disagreement, the final interpretation was determined by consensus. Clinical presentation and subsequent clinical course were reviewed to ensure consistency with the imaging findings; however, these factors were not used as primary diagnostic criteria.

At our institution, MRI is routinely performed in adolescent athletes with acute sports-related groin pain when radiographs are inconclusive, in order to evaluate a broad range of conditions that may be occult on plain radiographs, including muscle injury, stress fracture, apophyseal injury, and other traction-related pathologies. This institutional practice enables consistent assessment of radiographically occult injuries and minimizes the risk of selection bias in case identification. MRI was performed within 5 days of injury in all included cases.

### Data Collection

The following variables were extracted from medical records:

- Age and sex
- Skeletal maturity (open or closed physes)
- Primary sport
- Injury mechanism
- Ability to continue sports activity immediately after injury
- Treatment modality
- Time to return to sport

Return to sport was defined as the time point at which full return to competitive activity was permitted by the treating physician.

### Statistical Analysis

Only descriptive statistics were used. Continuous variables are presented as mean ± standard deviation, and categorical variables as counts and percentages.

### Ethics approval

This study was conducted in accordance with the principles of the Declaration of Helsinki. Approval was obtained from the institutional ethics committee prior to study initiation. Because of the retrospective nature of the study using existing clinical data, informed consent was obtained through an opt-out process, as approved by the ethics committee.

### Funding

The authors received no financial support for the research, authorship, and/or publication of this article.

## Results

Subperiosteal hematoma of the ilium was identified in 20 of 164 patients (12.2%) evaluated under the institutional MRI workflow for acute sports-related groin pain. The mean age of the 20 patients was 13.9 ± 1.5 years, and 19 patients (95%) were male. Open physes were observed in 18 patients (90%) (Table 1A).

**Table 1.** Demographic Characteristics, Sports Background, Injury Mechanism, and Clinical Course of Patients With Subperiosteal Hematoma of the Ilium (n = 20)

A. Demographic Characteristics
| Variable | Data |
| --- | --- |
| Age, years | 13.9 ± 1.5 |
| Male sex, n (%) | 19 (95) |
| Open growth plates, n (%) | 18 (90) |

B. Primary Sport and Injury Mechanism
| Variable | n (%) |
| --- | --- |
| **Primary sport** |  |
| Soccer | 13 (65) |
| Other sports* | 7 (35) |
| **Injury mechanism** |  |
| Kicking | 10 (50) |
| Sprinting | 4 (20) |
| Sudden deceleration | 3 (15) |
| Contact | 3 (15) |

\*Other sports included rugby (n=2), track and field (n=2), baseball (n=2), and judo (n=1).

C. Functional Severity and Clinical Course
| Variable | Data |
| --- | --- |
| Unable to continue sports activity, n (%) | 17 (85) |
| Conservative management, n (%) | 20 (100) |
| Time to return to sports, weeks | 4.5 ± 2.2 |

Soccer was the most common sport, accounting for 13 cases (65%), followed by rugby, track and field, and baseball, with 2 cases each. The mechanisms of injury included ball-kicking–related motion in 10 cases (50%), sprinting in 4 cases (20%), sudden deceleration in 3 cases (15%), and player-to-player contact in 3 cases (15%) (Table 1B).

Seventeen patients (85%) were unable to continue sports activity immediately after the injury. All patients were treated conservatively, and the mean time to return to sport was 4.5 ± 2.2 weeks (Table 1C).

## Discussion

This case series demonstrates a consistent clinical presentation characterized by (1) predominance in adolescent male soccer players, (2) frequent occurrence during kicking motions, (3) inability to continue sports activity immediately after injury, and (4) relatively rapid return to sport following conservative treatment.

In this consecutively evaluated clinical cohort of adolescent athletes presenting with acute sports-related groin pain under a standardized institutional imaging workflow, subperiosteal hematoma of the ilium was identified in 20 of 164 patients. Because this condition is typically occult on plain radiographs but clearly visualized on MRI, it may be overlooked in routine clinical evaluation [4–6, 8].

Importantly, this proportion reflects the relative frequency within this MRI-evaluated cohort and should not be interpreted as population incidence. This study suggests that subperiosteal hematoma of the ilium represents a reproducible traction injury pattern that becomes apparent only when MRI is incorporated into the diagnostic workflow for radiograph-negative groin pain [4, 5].

By analyzing a relatively homogeneous group of patients, this study demonstrated consistent patterns in patient demographics, injury mechanisms, and clinical course, including male predominance, a high prevalence of skeletal immaturity, concentration in soccer players, a strong association with kicking and sprinting motions, and a favorable return to sport with conservative management. These findings may assist clinicians in recognizing this entity when evaluating acute sports-related groin pain in adolescent athletes.

The injury mechanism observed in this cohort suggests traction stress applied to the periosteum of the iliac fossa during forceful hip movements such as kicking and sprinting. The predominance of adolescent soccer players and the association with these motions support this interpretation. This condition should be understood within the broader spectrum of apophyseal and traction-related injuries that occur during skeletal immaturity, rather than as an isolated hematoma. Similar to apophyseal avulsion injuries, the pathology arises from concentration of traction forces at anatomically vulnerable bone–periosteum interfaces characteristic of the growing skeleton [1, 10– 13].

However, MRI can demonstrate subperiosteal fluid collection, periosteal stripping, and surrounding reactive changes [4–8]. Recognizing this condition as an injury that may be occult on radiographs but detectable on MRI is important for avoiding diagnostic delay.

The differential diagnosis includes muscle strain, bone contusion, stress fracture, and apophyseal avulsion injury, and careful assessment of both imaging findings and the mechanism of injury can aid in differentiation [1–5].

Regarding treatment, all patients in this study were successfully managed conservatively and were able to return to sport within a relatively short period (mean, 4.5 weeks). The periosteum is a well-vascularized tissue with high healing potential, and when structural continuity is preserved, spontaneous recovery can be expected [10–12]. In the absence of structural instability or neurovascular compromise, activity modification and staged rehabilitation appear to be effective management strategies.

This study has several limitations. First, its retrospective single-center design and the absence of a control group limit the strength of causal inferences. Second, standardized imaging criteria and severity classification for this condition are not yet established. Third, long-term outcomes and recurrence rates were not fully evaluated. In addition, because MRI was frequently utilized in our institution when clinically indicated, the observed frequency may not be generalizable to settings where MRI is less readily performed. Therefore, the frequency observed in this study should not be interpreted as representing the true incidence in the general population. Future multicenter prospective studies are warranted to establish diagnostic criteria and optimal return-to-sport protocols.

Overall, subperiosteal hematoma of the ilium should be understood not as an incidental entity but as a form of traction injury associated with sports activity in adolescent athletes with growth-related anatomical characteristics [1, 10–13]. The absence of a standardized MRI grading system for this condition also limited assessment of imaging severity in relation to clinical recovery.

### Clinical Implications

In adolescent male athletes presenting with acute groin pain during kicking or sprinting and inability to continue play despite unremarkable radiographs, subperiosteal hematoma of the ilium related to iliacus traction should be considered. Early MRI evaluation should be considered in skeletally immature athletes presenting with acute groin pain, normal radiographs, and injury mechanisms involving kicking or sprinting.

## Conclusion

Awareness of this characteristic traction injury pattern may improve diagnostic accuracy in adolescent athletes presenting with acute groin pain and unremarkable radiographs.

Recognizing subperiosteal hematoma of the ilium as part of the spectrum of traction-related injuries during skeletal immaturity can help clinicians avoid misdiagnosis and facilitate appropriate management.

## Data Availability

All data produced in the present study are available upon reasonable request to the authors

---

**Figure 1.**
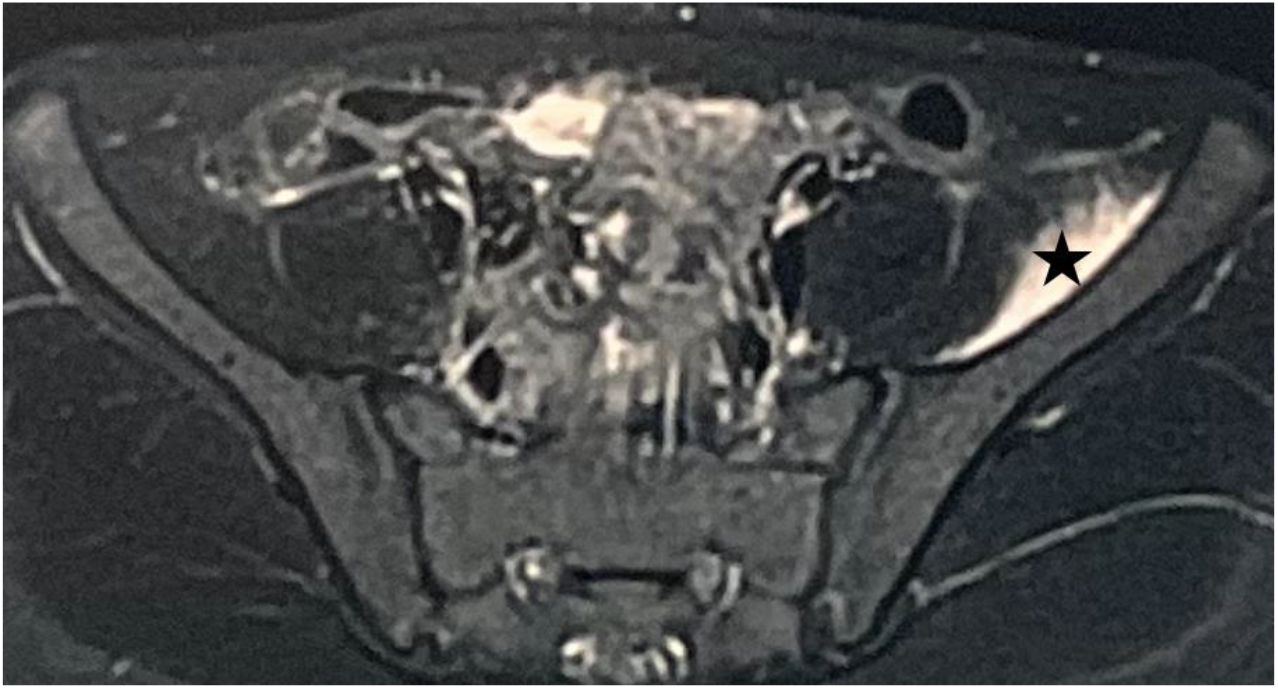
Axial T2-weighted fat-suppressed MRI of the pelvis obtained at initial presentation in an adolescent athlete with acute sports-related groin pain. A crescent-shaped hyperintense lesion (★) is seen along the inner cortex of the iliac fossa at the iliacus muscle attachment, indicating periosteal elevation with subperiosteal fluid accumulation. This lesion is typically undetectable on plain radiographs.

## Notes

**Conflict of Interest** The authors declare that they have no conflict of interest.

### Competing Interest Statement

The authors have declared no competing interest.

### Funding Statement

None.

### Author Declarations

The Ethics Committee of Ashiya Central Hospital gave ethical approval for this work (approval number: 7-ACH-11-2).

